# Data Extraction from Free-Text Stroke CT Reports Using GPT-4o and Llama-3.3-70B: The Impact of Annotation Guidelines

**DOI:** 10.1101/2025.01.22.25320938

**Authors:** Jonas Wihl, Enrike Rosenkranz, Severin Schramm, Cornelius Berberich, Michael Griessmair, Piotr Woźnicki, Francisco Pinto, Sebastian Ziegelmayer, Lisa C. Adams, Keno K. Bressem, Jan S. Kirschke, Claus Zimmer, Benedikt Wiestler, Dennis Hedderich, Su Hwan Kim

## Abstract

**Purpose:** To evaluate the performance of LLMs in extracting data from stroke CT reports in the presence and absence of an annotation guideline.

**Methods:** In this study, performance of GPT-4o and Llama-3.3-70B in extracting ten imaging findings from stroke CT reports was assessed in two datasets from a single academic stroke center. Dataset A (n = 200) was a stratified cohort including various pathological findings, whereas Dataset B (n = 100) was a consecutive cohort. Initially, an annotation guideline providing clear data extraction instructions was designed based on a review of cases with inter-annotator disagreements in dataset A. For each LLM, data extraction was performed under two conditions – with the annotation guideline included in the prompt and without it. Queries for both LLMs were run with a temperature setting of 0. For GPT-4o, additional queries with a temperature of 1 were performed.

**Results:** GPT-4o consistently demonstrated superior performance over Llama-3.3-70B under identical conditions, with micro-averaged precision ranging from 0.83 to 0.95 for GPT-4o and from 0.65 to 0.86 for Llama-3.3-70B. Across both models and both datasets, incorporating the annotation guideline into the LLM input resulted in higher precision rates, while recall rates largely remained stable. In dataset B, precision of GPT-4o and Llama-3-70B improved from 0.83 to 0.95 and from 0.87 to 0.94, respectively. The greatest increase in precision on a variable-level was seen in infarct demarcation (0.59 to 1.00) and subdural hematoma (0.67 to 1.00). Overall classification performance with and without annotation guideline was significantly different in five out of six conditions (e.g. dataset B/Llama-3.3/temp=0: p = 0.001).

**Conclusion:** Our results demonstrate the potential of GPT-4o and Llama-3.3-70B in extracting imaging findings from stroke CT reports, with GPT-4o steadily exceeding the performance of Llama-3-70B. We further provide evidence that well-defined annotation guidelines can enhance LLM data extraction accuracy.

## Introduction

CT imaging in suspected acute stroke, including non-enhanced CT (NECT), CT-angiography (CTA), and CT perfusion (CTP), provides critical insights into the type of stroke (ischemic or hemorrhagic), the presence of vessel occlusions, and the extent of brain damage. The imaging findings, which are recorded in radiological reports, are pivotal in determining eligibility for intravenous thrombolysis or mechanical thrombectomy. Importantly, the data contained in these CT reports holds enormous value beyond its utility in clinical decision-making, enabling various studies on epidemiology (1), pathophysiology (2), treatment efficacy (3,4), and patient outcomes (5,6). Key data variables can further be utilized as labels for training machine learning algorithms for tasks such as detecting large vessel occlusion (7), automatic evaluation of ASPECTS (8), infarct segmentation (9,10), and lesion classification (11). Imaging findings also play a crucial role in national stroke registries aiming to monitor and improve quality of stroke care (12,13).

Yet, given that most radiology reports still consist of prose and lack standardized terminology, large-scale analysis of textual descriptions of imaging findings previously necessitated labor-intensive manual annotations by clinical experts, thereby limiting scalability (14). Natural language processing (NLP) systems based on machine learning have shown promising results in automating information extraction from radiology reports, but were limited by the scarcity of annotated training data as well as the variability and ambiguity of reports (15–17).

Recently, large language models (LLMs) have demonstrated great potential in overcoming these limitations. LLMs are artificial intelligence (AI) systems trained on vast datasets and capable of performing various natural language tasks such as text classification, summarization and generation (18). In radiology, LLMs have shown significant promise in tasks such as report generation (19,20), report translation (21), differential diagnosis (22,23), and patient education (24). The performance of LLMs in data extraction has been evaluated in different imaging modalities ranging from X-ray to interventional angiography (25–29), and studies most frequently focused on chest and head examinations (30). Notably, both proprietary (25,27,28) and open-source LLMs (26,29) have been assessed, with open-source models offering the advantage of local data processing as an additional layer of safety to protect patient data privacy. In models of both categories, high accuracy levels of more than 90% of correctly extracted parameters have been reported, demonstrating their potential utility.

However, methodical inconsistencies in these types of studies pose challenges. For instance, many studies relied on manual annotations performed by only a single annotator (27,28,31), making the gold standard prone to subjective bias and human error. In addition, a scoping review by Reichenpfader et al points out that only 9% (3/34) of studies reported annotation guidelines (30), unveiling a frequent lack of standardization and transparency in the annotation process. Moreover, many studies modeled findings in radiology reports as simple binary variables, which fails to capture the nuanced levels of diagnostic uncertainty expressed in the textual descriptions (26,30,32).

Against this background, the aim of this study was to evaluate the performance of LLMs in data extraction from head CT reports in suspected acute stroke in the presence and absence of a comprehensive annotation guideline.

## Methods

This retrospective study was approved by the Institutional Review Board of the Technical University of Munich (TUM) and the need for informed consent was waived.

### Datasets

This study employed two different datasets from a single German academic institution with a comprehensive stroke center. Both datasets included patients who underwent a CT examination in suspected acute stroke, using protocols that featured either unenhanced CT with CT angiography only, or an additional CT perfusion. Reports were available in German language.

Dataset A (n = 200) was a stratified cohort comprising five purposively sampled subgroups with the following imaging findings each (exam dates ranging from 14 June 2022 to 14 July 2024): ischemic stroke of the anterior circulation (n = 40), ischemic stroke of the posterior circulation (n = 40), extracranial pathology (e.g. carotid stenosis or arterial dissection) (n = 40), intracranial hemorrhage (n = 40), and miscellaneous pathologies (n = 40). Covering a variety of pathological findings, this dataset served as the basis for creating a comprehensive annotation guideline. Dataset B (n = 100) contained a chronologically collected, consecutive cohort between 1 August 2024 and 14 September 2024.

### Data Extraction Parameters

A template with the following ten imaging findings was created and represented in JavaScript Object Notation (JSON) format (Figure 1): intracerebral hemorrhage (ICH), epidural hemorrhage (EDH), subdural hemorrhage (SDH), subarachnoid hemorrhage (SAH), infarct demarcation, vascular occlusion, vascular stenosis, aneurysm, dissection, and perfusion deficit.

**Figure 1:**
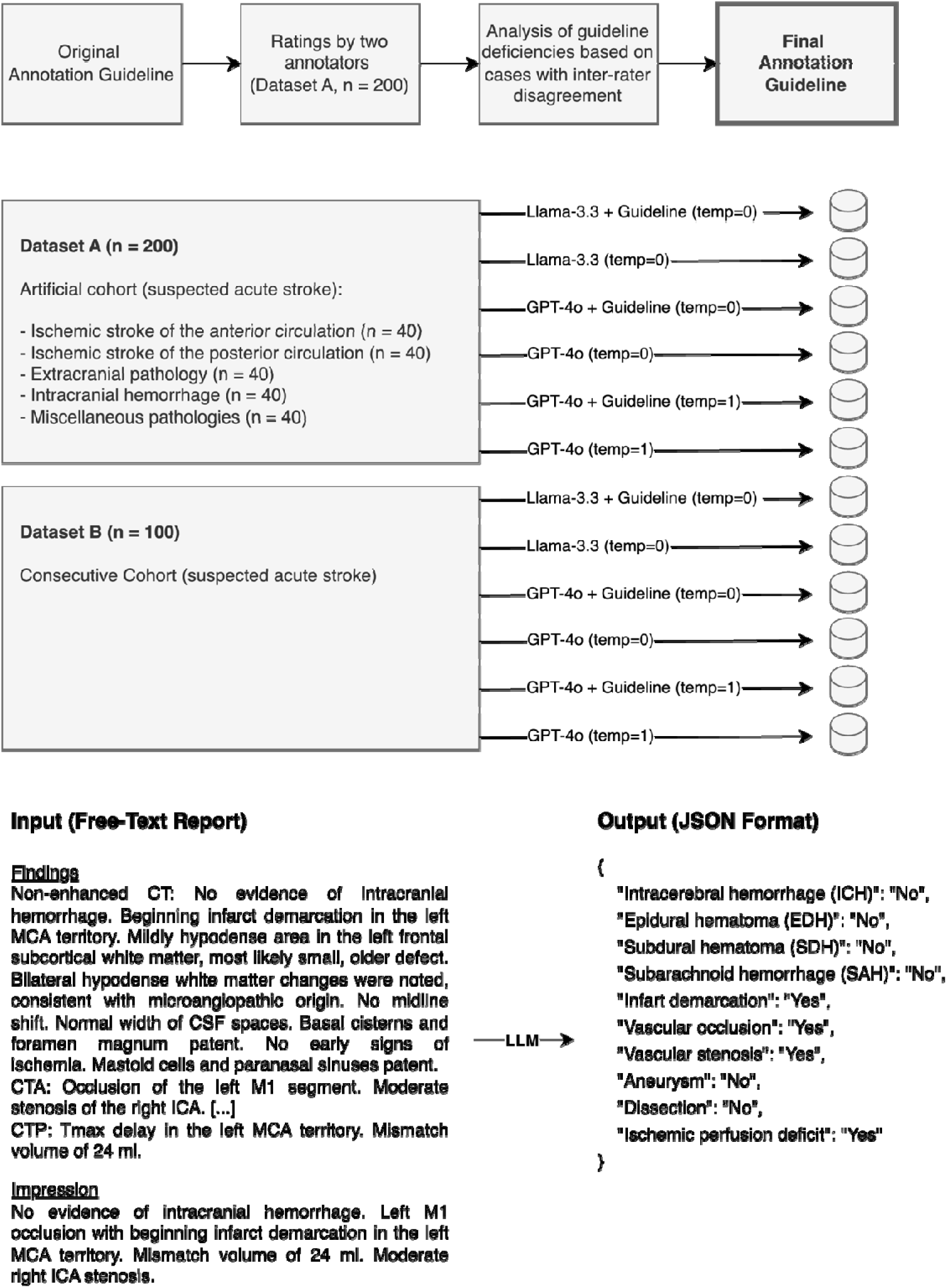
Study Design. Initially, two raters annotated dataset A using a preliminary annotation guideline with a few general instructions. Based on the guideline deficiencies uncovered based on a review of cases with inter-rater disagreement, an addendum was appended to the original document, forming the final annotation guideline. The data extraction performance of GPT-4o and Llama-3.3-70B with and without annotation guideline was evaluated in dataset A and additionally in another dataset (dataset B) that was not used to formulate the annotation guideline. At the bottom, a fictional CT report in English is shown along with the data parameters extracted from it in JSON format to illustrate the methodology.

### Manual Annotations and Annotation Guideline

A prototypic user interface (provided by Smart Reporting GmbH, Munich, Germany), was used to perform manual data entries. One radiology resident with two years of dedicated neuroradiology experience (SHK) and one fourth-year medical student (JW) independently annotated dataset A (n = 200). A brief annotation guideline with general instructions (e.g. handling of missing data) defined by SHK was followed by both annotators. During the annotation process, ambiguous instances were identified and recorded by the raters.

SHK and DMH (a board-certified neuroradiologist with 10 years of experience) reviewed cases with inter-rater disagreement and, added a detailed addendum to the original annotation guideline addressing the identified edge cases. Manual annotations of dataset A were revised according to the final guideline. Manual annotations of dataset B were equally conducted by SHK and JW according to the final guideline, and cases of inter-rater disagreement were resolved by DMH.

Following the closed-world assumption, findings not mentioned in the report were considered absent. Findings with uncertainty descriptors (“possible”, “DDx”, etc.) indicating no clear positive or negative tendency were labeled as “unknown” by annotators and omitted from the LLM data extraction analysis.

### LLM Infrastructure

GPT-4o (‘gpt-4o-2024-08-06’) by OpenAI (33) and Llama-3.3-70B (‘Llama-3.3-70B-Instruct’) by Meta (34) were chosen as representative state-of-the-art proprietary and open-source LLM each at the time of the study. GPT-4o was accessed via OpenAI’s application programming interface (API) (https://platform.openai.com/docs/models#gpt-4o). Llama-3.3-70B was deployed in a local environment utilizing the Python library “llama-cpp-python”, which provides compressed, less memory-intensive LLM instances (‘quantization’). A quantization factor of Q4_K_M was chosen to allow full GPU offloading. A single NVIDIA Quadro RTX 8000 with 48 GB of video memory was used for local inferences.

For both GPT-4o and Llama-3.3-70B, the model temperature was set to 0.0 to ensure deterministic results. To explore the impact of temperature settings on data extraction performance, GPT-4o queries were additionally run with the default temperature setting of 1. Our scripts for executing both models are publicly available in our repository at: https://github.com/shk03/stroke_llm_data_extraction.

### LLM Queries

For both models, queries were performed with and without annotation guidelines each. The base prompt was defined as follows (translated from German to English):

“*Extract the information provided in the radiological report in the format of a JSON file. Each of the ten parameters should be evaluated as ‘Yes’ or ‘No.’ Findings that are not mentioned are considered absent and should be evaluated as ‘No.’*

*Please take the following guidelines into account:*

*{annotation_guidelines}*

*The JSON file should have the following structure:*

*{json_schema}*

*Here is the report from which the information should be extracted:*

*{report_text}*”

Queries with a temperature of 0 were conducted only once, assuming that this setting would lead to almost fully deterministic results. In contrast, GPT-4o queries with a temperature of 1 were repeated three times each to account for probabilistic variations. Execution times for LLM queries were recorded.

### GPT-4o Performance in Diagnostic Certainty Assessment

In an additional experiment, the ability of GPT-4o to correctly evaluate diagnostic uncertainty of report content was evaluated in dataset B, using the default temperature setting of 1. GPT-4o was instructed to classify the ten parameters into one of the following five categories: ‘certainly absent’, ‘unlikely’, ‘possible’, ‘likely’, ‘certainly present’. Accuracy was rated against manual annotations by a single annotator (JW).

### Analysis

Statistical analyses and data visualizations were performed using the libraries Pandas and Matplotlib in Python (Version 3.11.8). To calculate accuracy metrics for GPT-4o queries with a temperature of 1, the mode of the labels across three repetitions was used. The extent of probabilistic variation was quantified as the percentage of cases producing consistent results across three repetitions. For both models, precision (= positive predictive value), recall (= sensitivity), and F1 (= harmonic mean of precision and recall) were reported. Aggregated metrics across all extracted parameters were computed using micro-averaging, which consolidates true positives, false positives, and false negatives globally. Confidence intervals for precision and recall were determined using the Wilson score method (35). The resulting lower and upper bounds were used to approximate the confidence interval of the F1 score. Overall classification performances were compared between groups with and without annotation guideline using the McNemar’s test. A p-value of < 0.05 was considered statistically significant. Correction for multiple testing was not performed, given the exploratory nature of the study.

## Results

### Cohort Overview

An overview of the patient cohorts is presented in Table 1. Patients in both datasets had a median age of 79 (A: IQR of 72 – 85, B: IQR of 65 - 84), and an equal sex distribution (50.0 and 51.0% females each). Due to its purposive selection, dataset A exhibited a higher proportion of pathological findings than the consecutive dataset B, with particularly high occurrences of vascular occlusions (A: 43.0%, B: 21.0%), stenoses (A: 38.5%, B: 26.0%) and ischemic perfusion deficits (A: 42.5%, B: 25.0%). Intracranial hemorrhages were relatively rare, with intracerebral hemorrhages being most common in both datasets (A: 8.5%, B: 4.0%). Similarly, aneurysms (A: 4.5%, B: 4.0%) and arterial dissections (A: 1.5%, B: 1.0%) were found only in rare occasions.

**Table 1:**
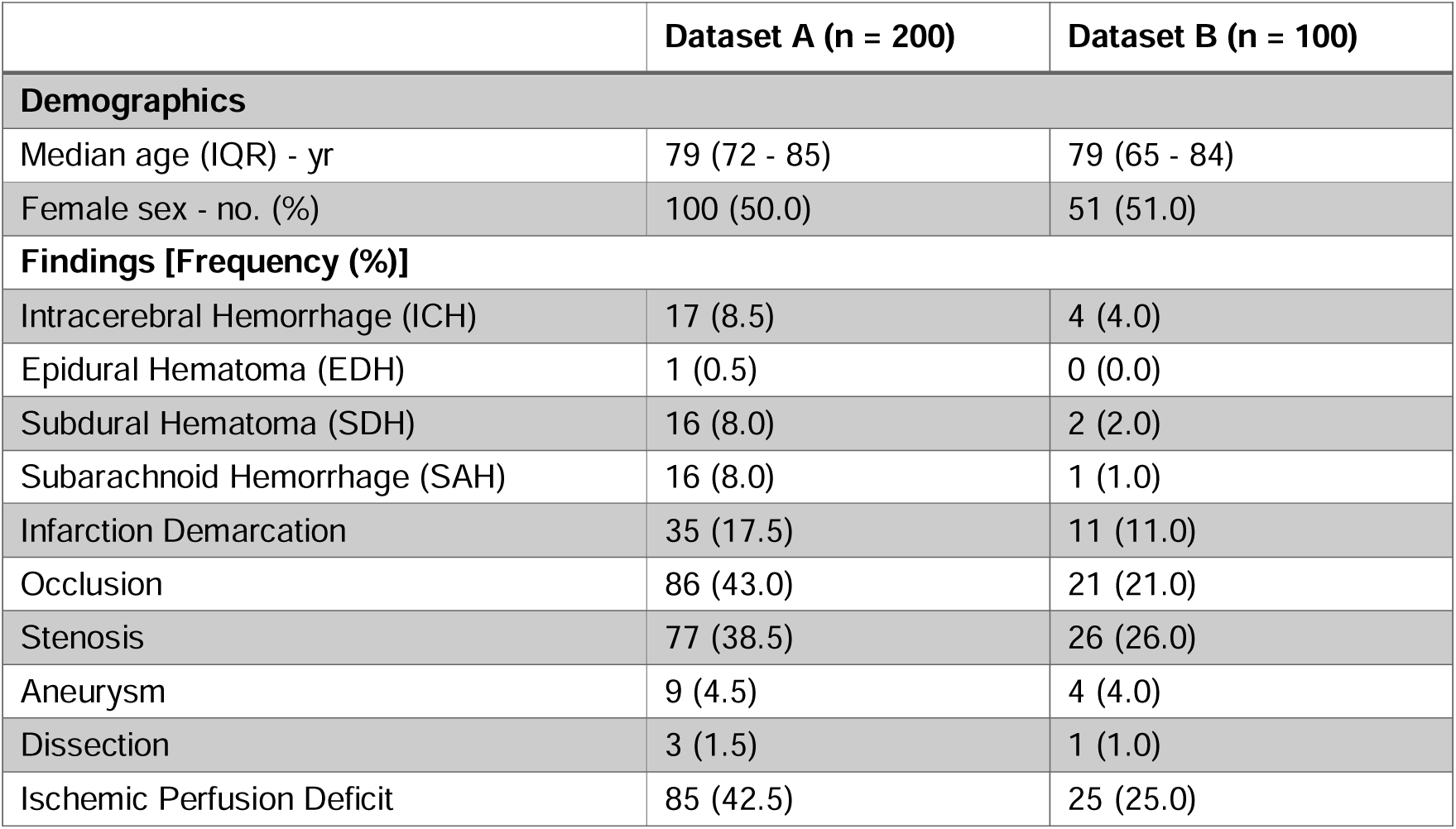
Cohort Overview. Dataset A was a stratified cohort with 40 cases of the following five subgroups each: ischemic stroke of the anterior circulation, ischemic stroke of the posterior circulation, extracranial pathology, intracranial hemorrhage, miscellaneous pathologies. Dataset B was a consecutive cohort of cases with suspected acute stroke.

|  | Dataset A (n = 200) | Dataset B (n = 100) |
| --- | --- | --- |
| **Demographics** |  |  |
| Median age (IQR) - yr | 79 (72 - 85) | 79 (65 - 84) |
| Female sex - no. (%) | 100 (50.0) | 51 (51.0) |
| **Findings [Frequency (%)]** |  |  |
| Intracerebral Hemorrhage (ICH) | 17 (8.5) | 4 (4.0) |
| Epidural Hematoma (EDH) | 1 (0.5) | 0 (0.0) |
| Subdural Hematoma (SDH) | 16 (8.0) | 2 (2.0) |
| Subarachnoid Hemorrhage (SAH) | 16 (8.0) | 1 (1.0) |
| Infarction Demarcation | 35 (17.5) | 11 (11.0) |
| Occlusion | 86 (43.0) | 21 (21.0) |
| Stenosis | 77 (38.5) | 26 (26.0) |
| Aneurysm | 9 (4.5) | 4 (4.0) |
| Dissection | 3 (1.5) | 1 (1.0) |
| Ischemic Perfusion Deficit | 85 (42.5) | 25 (25.0) |

### Annotation Guidelines

The content of the original and final annotation guideline is presented in Table 2. The original annotation guideline was used by the two raters during initial annotation of dataset A, and contained general instructions on handling purely descriptive image findings, indicators of diagnostic uncertainty, and contradictions within the report.

**Table 2:** Annotation Guideline.

| Version | Type | Instruction |
| --- | --- | --- |
| Original | General | **No Interpretation of Findings**<br>A finding is only deemed present if it is explicitly mentioned.<br>Examples:<br>- Hypodensity in the parenchyma is not equivalent to infarct demarcation.<br>- "Calcifications" or "plaques" are not equivalent to stenosis.<br>- "Intraluminal thrombus" is not equivalent to stenosis.<br>- "Loss of corticomedullary differentiation" is not equivalent to infarct demarcation.<br>- "Contrast agent discontinuation," "vessel discontinuation," or "missing vascular contrast" is equivalent to vessel occlusion.<br>- If the ASPECTS score is below 10, it is classified as infarct demarcation. |
| Original | General | **Diagnostic Certainty**<br>- Findings with a positive tendency (e.g., "most likely," "suspicion of," "probable") are classified as "Yes".<br>- Findings with a negative tendency (e.g., "unlikely," "secondary consideration") are classified as "No". |
| Original | General | **Contradictions**<br>- In case of contradictions between the "Findings" and "Impression" sections, the "Impression" is considered decisive, and the finding is classified accordingly. |
| Addendum | General | **Exclusion of Previous Findings and Medical History**<br>- Previous findings that are not present anymore are not considered.<br>- Previous medical history is not considered. |
| Addendum | General | **Location of Findings**<br>- A finding shall be considered present if at least one location is affected. For example, "stenosis" shall be evaluated as "Yes" if a stenosis is described in at least one location. |
| Addendum | Specific | **Infarct Demarcation**<br>- "Infarct demarcation" encompasses only acute changes. Chronic infarcts or post-ischemic changes shall not be counted. |
| Addendum | Specific | **Aneurysms**<br>- "Aneurysm" refers exclusively to true intracranial aneurysms. Aortic aneurysms or pseudoaneurysms shall not be classified as aneurysms. |
| Addendum | Specific | **Perfusion Deficit** |
|  |  | - A Tmax delay shall be considered a perfusion deficit.<br>- Perfusion deficits do not require a mismatch.<br>- Only perfusion deficits attributed to ischemia in the report shall be counted. Perfusion deficits due to artifacts or other causes shall not be counted. |
| Addendum | Specific | **Stenosis / Occlusion**<br>- In cases where a high-grade stenosis or occlusion is indicated ("stenosis/occlusion"), "stenosis" shall be evaluated as present and "occlusion" as "No."<br>- Vascular irregularities shall not be classified as stenosis.<br>- Vascular occlusion refers exclusively to arterial occlusions (excluding sinus or cerebral venous thrombosis).<br>- Pre-existing occlusions shall also be classified as vascular occlusion. |

Annotators agreed in 96.0% (1920/2000) of data points (Cohens’ kappa κ = 0.852). A thorough review of cases with inter-annotator disagreement (4.0%; 80/2,000) revealed 1.3% (26/2,000) of discrepancies originating from unclear guideline instructions, as opposed to 2.7% (54/2,000) of cases resulting from careless mistakes. To address the identified guideline deficiencies, an addendum was created which contained both general instructions (e.g. handling mentions of previous findings) and directions on classifying edge cases for specific parameters (e.g. not counting pseudoaneurysms as aneurysms). Using a few-shot prompting approach, some instructions in the annotation guideline included one or more example expression, along with the correct label. The final annotation guideline containing the original instructions and addendum was provided to GPT-4o and Llama-3.3-70B.

### Model Performance

1.7% (34/2000) and 1.4% (14/1000) of data points were excluded from dataset A and dataset B respectively, as the report text indicated diagnostic uncertainty without a clear positive or negative tendency (expressions such as “possible”, “DDx”).

Overall, GPT-4o consistently demonstrated superior performance over Llama-3.3-70B under identical conditions, with micro-averaged precision (= positive predictive value) ranging from 0.86 to 0.95 for GPT-4o and from 0.65 to 0.86 for Llama-3.3-70B. In Dataset B, GPT-4o and Llama-3.3-70B (temperature = 0) yielded a precision of 0.95 and 0.74 each in the presence of the annotation guideline, while both exhibited equal recall values (= sensitivity; both 0.98). Across all conditions, higher precision rates were observed when the annotation guideline was available.

The precision of GPT-4o (temperature = 1) improved from 0.87 to 0.94 in dataset A (p < 0.001) and from 0.83 to 0.95 in dataset B (p = 0.006). When using a temperature of 0, GPT-4o’s precision increase from 0.86 to 0.95 in dataset A (p < 0.001) and from 0.86 to 0.93 in dataset B, although the difference under these conditions was not significant (p = 0.390). Similarly, the precision of Llama-3.3 (temperature = 0) rose from 0.78 to 0.86 (p < 0.001) in dataset A and from 0.87 to 0.94 in dataset B (p = 0.001). In contrast, recall rates largely remained stable, with values ranging from 0.98 to 0.99 in all conditions (Figure 2).

**Figure 2:**
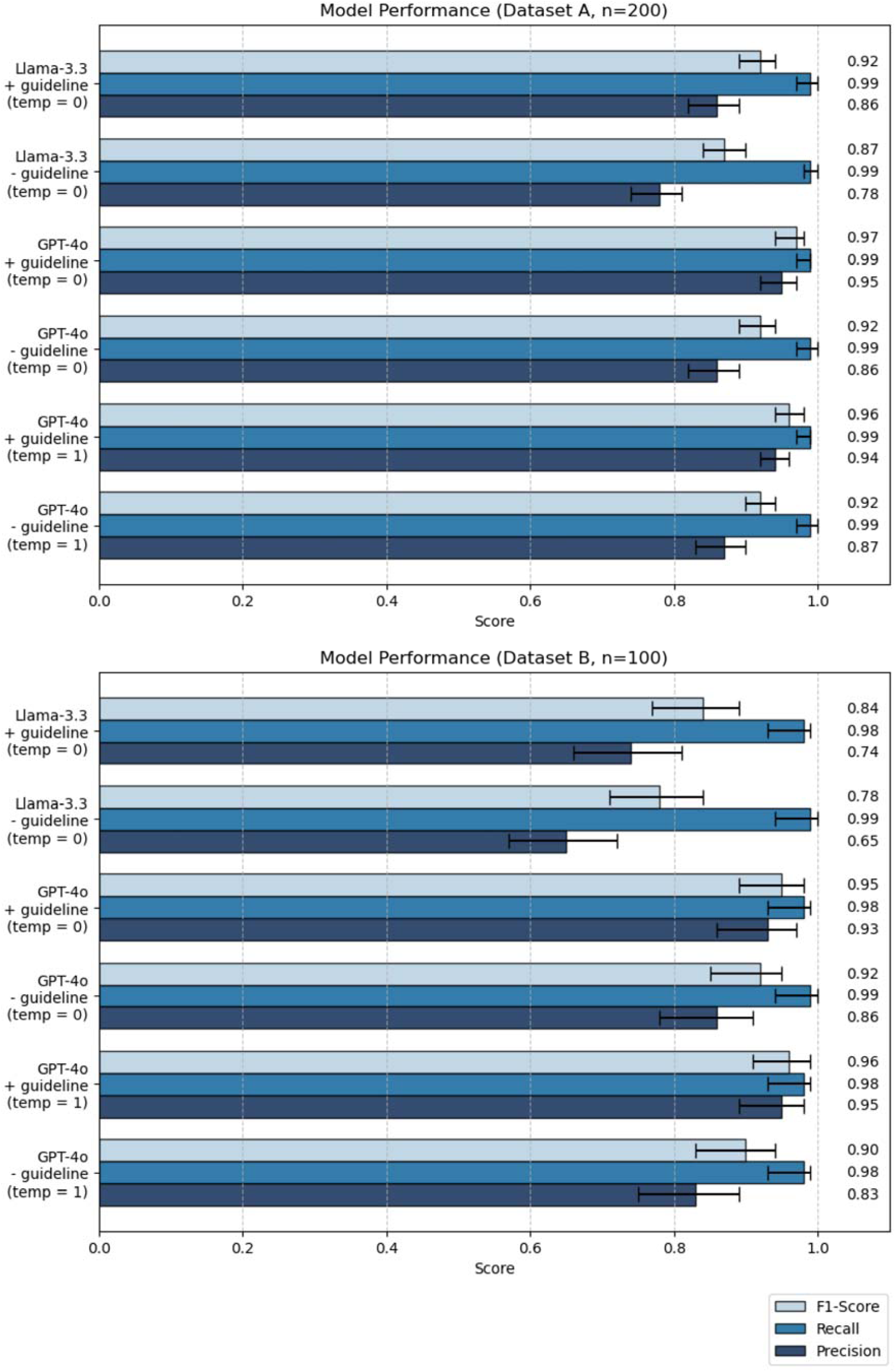
Data Extraction Performance of GPT-4o and Llama-3.3-70B across all parameters. Metrics for GPT-4o queries with a temperature setting of 1 were calculated based on the mode across three repetitions, whereas remaining queries were run only once (with a temperature setting of 0.0). Error bars indicate 95% confidence intervals. 1.7% (34/2000) and 1.4% (14/1000) of data points were excluded from dataset A and dataset B each, as the report text indicated diagnostic uncertainty without a clear positive or negative tendency (expressions such as “possible”, “DDx”). Precision: positive predictive value. Recall: sensitivity. F1-Score: harmonic mean of precision and recall.

Temperature settings of GPT-4o had only a minor impact on data extraction performance when the remaining conditions were equal. The largest difference in precision was seen in dataset B in the absence of guidelines, where an increase of the temperature resulted in a small drop in precision from 0.86 (temp = 0) to 0.83 (temp = 1).

Detailed variable-level metrics of GPT-4o in dataset B (temperature = 1) are presented in Table 3. Metrics varied widely, particularly in findings with low prevalence in the given dataset (e.g. 0 cases with true epidural hematoma, 1 case with subarachnoid hemorrhage). The greatest increase in precision through the guideline adoption was seen in infarct demarcation (0.59 to 1.00), subdural hematoma (0.67 to 1.00), and vascular stenosis (0.84 to 0.96). Exemplary cases where the annotation guideline influenced the LLM ratings are shown in Table 4. Granular variable-level metrics of remaining groups are provided in Supplement 1-5.

**Table 3:**
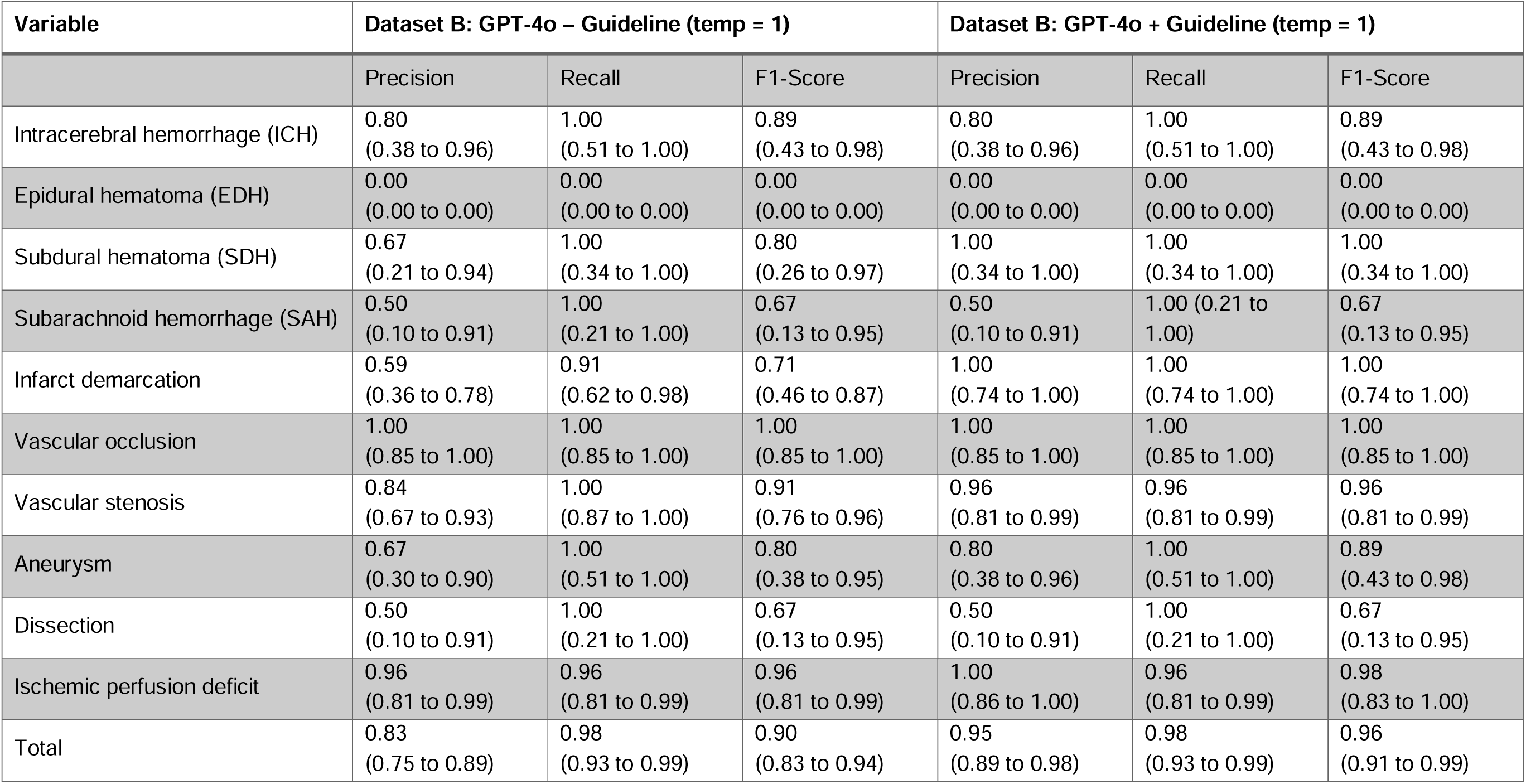
Data extraction performance of GPT-4o (temperature = 1) with and without annotation guideline in dataset B (n = 100). 1.4% (14/1000) of data points were excluded, as the report text indicated diagnostic uncertainty without a clear positive or negative tendency (expressions such as “possible”, “DDx”). Metrics for GPT-4o were calculated based on the mode across three repetitions.

| Variable | Dataset B: GPT-4o – Guideline (temp = 1) |  |  | Dataset B: GPT-4o + Guideline (temp = 1) |  |  |
| --- | --- | --- | --- | --- | --- | --- |
|  | Precision | Recall | F1-Score | Precision | Recall | F1-Score |
| Intracerebral hemorrhage (ICH) | 0.80<br>(0.38 to 0.96) | 1.00<br>(0.51 to 1.00) | 0.89<br>(0.43 to 0.98) | 0.80<br>(0.38 to 0.96) | 1.00<br>(0.51 to 1.00) | 0.89<br>(0.43 to 0.98) |
| Epidural hematoma (EDH) | 0.00<br>(0.00 to 0.00) | 0.00<br>(0.00 to 0.00) | 0.00<br>(0.00 to 0.00) | 0.00<br>(0.00 to 0.00) | 0.00<br>(0.00 to 0.00) | 0.00<br>(0.00 to 0.00) |
| Subdural hematoma (SDH) | 0.67<br>(0.21 to 0.94) | 1.00<br>(0.34 to 1.00) | 0.80<br>(0.26 to 0.97) | 1.00<br>(0.34 to 1.00) | 1.00<br>(0.34 to 1.00) | 1.00<br>(0.34 to 1.00) |
| Subarachnoid hemorrhage (SAH) | 0.50<br>(0.10 to 0.91) | 1.00<br>(0.21 to 1.00) | 0.67<br>(0.13 to 0.95) | 0.50<br>(0.10 to 0.91) | 1.00 (0.21 to 1.00) | 0.67<br>(0.13 to 0.95) |
| Infarct demarcation | 0.59<br>(0.36 to 0.78) | 0.91<br>(0.62 to 0.98) | 0.71<br>(0.46 to 0.87) | 1.00<br>(0.74 to 1.00) | 1.00<br>(0.74 to 1.00) | 1.00<br>(0.74 to 1.00) |
| Vascular occlusion | 1.00<br>(0.85 to 1.00) | 1.00<br>(0.85 to 1.00) | 1.00<br>(0.85 to 1.00) | 1.00<br>(0.85 to 1.00) | 1.00<br>(0.85 to 1.00) | 1.00<br>(0.85 to 1.00) |
| Vascular stenosis | 0.84<br>(0.67 to 0.93) | 1.00<br>(0.87 to 1.00) | 0.91<br>(0.76 to 0.96) | 0.96<br>(0.81 to 0.99) | 0.96<br>(0.81 to 0.99) | 0.96<br>(0.81 to 0.99) |
| Aneurysm | 0.67<br>(0.30 to 0.90) | 1.00<br>(0.51 to 1.00) | 0.80<br>(0.38 to 0.95) | 0.80<br>(0.38 to 0.96) | 1.00<br>(0.51 to 1.00) | 0.89<br>(0.43 to 0.98) |
| Dissection | 0.50<br>(0.10 to 0.91) | 1.00<br>(0.21 to 1.00) | 0.67<br>(0.13 to 0.95) | 0.50<br>(0.10 to 0.91) | 1.00<br>(0.21 to 1.00) | 0.67<br>(0.13 to 0.95) |
| Ischemic perfusion deficit | 0.96<br>(0.81 to 0.99) | 0.96<br>(0.81 to 0.99) | 0.96<br>(0.81 to 0.99) | 1.00<br>(0.86 to 1.00) | 0.96<br>(0.81 to 0.99) | 0.98<br>(0.83 to 1.00) |
| Total | 0.83<br>(0.75 to 0.89) | 0.98<br>(0.93 to 0.99) | 0.90<br>(0.83 to 0.94) | 0.95<br>(0.89 to 0.98) | 0.98<br>(0.93 to 0.99) | 0.96<br>(0.91 to 0.99) |

**Table 4:** Exemplary data extraction cases influenced by the annotation guideline (GPT-4o, Dataset B, temp=1). Report excerpts were translated from German to English.

| Parameter | Report Excerpt | Relevant Guideline Instruction | Rating:<br>GPT-4o<br>– Guideline | Rating:<br>GPT-4o<br>+ Guideline | Correct<br>Rating |
| --- | --- | --- | --- | --- | --- |
| Subdural hematoma (SDH) | No evidence of hyperdensities suggestive of bleeding. S/p left frontal trepanation for subdural hematoma. | Previous findings that are not present anymore are not considered. | Yes | No | No |
| Stenosis | Calcified plaques at the carotid bulb bilaterally. | A finding is only deemed present if it is explicitly mentioned. "Calcifications" or "plaques" are not equivalent to stenosis. | Yes | No | No |
| Stenosis | Caliber irregularities of the left vertebral artery with calcified plaque in the V4 segment. Along the right vertebral artery, multiple calcified plaques are observed. | "Calcifications" or "plaques" are not equivalent to stenosis. Vascular irregularities are not considered stenosis. | Yes | No | No |
| Infarct demarcation | Old partial infarction of the right MCA territory. No early signs of ischemia, no evidence of intracranial hemorrhage. | "Infarct demarcation" encompasses only acute changes. Chronic infarcts or post-ischemic changes shall not be counted. | Yes | No | No |
| Infarct demarcation | Hypodense parenchymal defect area in the right occipital lobe. | A finding is only deemed present if it is explicitly mentioned. Hypodensity in the parenchyma is not equivalent to infarct demarcation. | Yes | No | No |
| Aneurysm | Bilateral ICA pseudoaneurysms. | "Aneurysm" refers exclusively to true intracranial aneurysms. Aortic aneurysms or pseudoaneurysms shall not be classified as aneurysms. | Yes | No | No |
| Ischemic perfusion deficit | Slight Tmax delay in the left high frontal and high parietal regions with partially increased CBF, DDx postictal. | Only perfusion deficits attributed to ischemia in the findings text shall be counted. Perfusion deficits due to artifacts or other causes shall not be counted. | Yes | No | No |

### Processing Time and Test-Retest Reliability

Mean processing times for GPT-4o (accessed via API) was 465.1 seconds per 100 reports, as compared to 1441.4 seconds per 100 reports for Llama-3.3-70B (operated on a local GPU). Mean time for manual annotations (measured in dataset B) was considerably longer than both models (9302.0 seconds per 100 reports). GPT-4o with a temperature setting of 1 featured a very high test-retest reliability, with identical ratings across three repetitions in 97.6% of data points.

### Diagnostic Certainty Assessment

GPT-4o accurately classified the diagnostic certainty level in 90.0% (900/1,000) of data points in dataset B. Yet, its performance was considerably lower in uncertain findings (categories ‘unlikely’, ‘possible’, ‘likely’), with only 35.0% (7/20) correct data points. In contrast, it performed markedly higher for certain findings (categories ‘definitely absent’, ‘definitely present’), achieving 91.1% accuracy (893/980).

## Discussion

In this study, we evaluated the performance of GPT-4o and Llama-3.3-70B in extracting data parameters from stroke CT reports with and without comprehensive annotation guidelines.

In summary, we demonstrate promising performance of GPT-4o and Llama-3.3-70B in extracting key imaging findings from stroke CT reports. Our results extend the findings from previous studies illustrating the utility of LLMs in extracting data parameters from mechanical thrombectomy reports (28,29), brain MRI reports (26), and more. Although GPT-4o invariably outperformed Llama-3.3-70B given the identical dataset and condition, Llama-3.3-70B showed great potential, with overall precision and recall scores of up to 0.86 and 0.99 each. This is in accordance with several recent studies highlighting that open-source models are rapidly catching up with state-of-the-art proprietary models in clinical tasks (36–38). However, numerous advantages of open-source models for clinical use have been pointed out by authors, including enhanced data privacy, greater control over updates and customization, transparency, and stronger community collaboration (29,39,40). Hence, it is reasonable to expect continued interest in and support for open-source models among the medical community, even though their local implementation demands great technical expertise and an advanced hardware infrastructure.

To explore the role of LLM temperature, we performed GPT-4o queries with two different settings (0 and 1) but observed only negligible differences in data extraction metrics. This is in alignment with a previous study on LLM-based information extraction from clinical trial publications reporting consistent performance in a temperature range of 0 – 1.5 (41). In general, temperature is considered a LLM hyperparameter influencing the randomness and creativity of model outputs. In medicine, it might be reasonable to avoid too high temperature levels that could lead to more frequent hallucinations (42).

Crucially, this study emphasizes the role of a comprehensive annotation guideline on LLM data extraction performance. In both models and both datasets evaluated, the inclusion of a detailed annotation guideline led to a substantial increase in precision, while retaining very high recall scores. The employed annotation guideline, which was equally adopted by the human annotators defining the reference standard, included detailed definitions of the individual data variables, along with instructions for specific edge cases. A more granular analysis on the variable level reveals that the improvement in data extraction metrics was primarily driven by a more precise and narrower definition of several key parameters including ‘infarct demarcation’ and ‘vascular stenosis’.

The guideline additionally provided directions on handling diagnostic uncertainty of findings, which is an inherent limitation of diagnostic interpretations in medical imaging. A wide variability in phrases conveying certainty levels in radiology reports has been reported previously (43,44), and some authors proposed the adoption of standardized certainty scales to improve clinical communication (45). While uncertainty of findings cannot be eliminated, the data annotation process required a categorization into a binary variable. To resolve this issue, our annotation guideline specified that uncertain findings with a clear positive or negative tendency be classified as “Yes” or “No” respectively, though equivocal findings (e.g. “possible”, “DDx”) were manually excluded for the purpose of the analysis.

In a complementary experiment, we observed that GPT-4o displayed high accuracy in classifying findings on a 5-point certainty scale if findings were definitive (‘definitely present’, ‘definitely absent’) but struggled to correctly assign uncertain findings (‘likely’, ‘possible’, ‘unlikely’), suggesting a potential weakness.

It is worth noting that the annotation guideline in this study was defined based on a meticulous review of cases with inter-annotator disagreement in one of the two datasets. This approach uncovered numerous edge cases and rating ambiguities that had not been anticipated in advance. When applying LLM-based data extraction in real-world use cases, annotation guidelines should be carefully designed to reflect the intended downstream use of the extracted data. For example, a more restrictive definition of variables leading to higher precision might be sensible if the accurate identification of certainly positive cases is decisive (e.g. in a retrospective study with strict inclusion criteria), whereas higher sensitivity (recall) should be prioritized if the primary goal is not to miss any true positives (e.g. identifying critical incidents).

Despite the fast-paced advancement of LLM capabilities, data extraction from unstructured radiology reports is constrained by their lacking standardization of content and terminology. Classifications such as ASPECTS (Alberta Stroke Program Early CT Score) (46), that are frequently assessed in study settings, cannot be meaningfully analyzed if not routinely reported. Extracting the location of a finding is complicated by its variable description (e.g. in terms of adjacent structures, vascular territories, or brain lobes). Furthermore, findings that are not explicitly stated introduce another layer of ambiguity. In our study, both human annotators and LLMs generally operated under the “closed-world assumption”, whereby findings were presumed absent unless explicitly mentioned. Nevertheless, it is possible that in some cases, lacking mentions are indicative of findings missed by the radiologist. The impact of this ambiguity on clinical communication was exemplified in a survey study, where half of the referring clinicians believed the radiologist might not have evaluated a particular feature if not explicitly documented in the report (47).

### Limitations

Several limitations need to be acknowledged. First, the single-center nature of this study necessitates further validation to confirm the generalizability of our findings. Second, due to the relatively small sample size of the consecutive cohort and the low occurrence of some findings in the dataset, the variable-level analysis of data extraction metrics was underpowered. Third, this study utilized only German reports, and the influence of language on LLM performance has not been explicitly assessed. Finally, the performance of guideline-enhanced LLMs in Dataset A needs to be interpreted with caution, given that the annotation guideline was derived from ambiguous cases within the same dataset.

### Conclusion

Our results demonstrate the potential of GPT-4o and Llama-3-70B in extracting key image findings from stroke CT reports, with GPT-4o steadily exceeding the performance of Llama-3-70B. We further provide evidence that well-defined annotation guidelines can enhance LLM data extraction accuracy.

## Data Availability

The code for running the LLM queries is provided online at https://github.com/shk03/stroke_llm_data_extraction

