## Supplementary Tables for "Data Extraction from Free-Text Stroke CT Reports Using GPT-4o and Llama-3.3-70B: The Impact of Annotation Guidelines"

| Variable | Dataset A: GPT-4o – Guideline (temp = 1) | | | Dataset A: GPT-4o + Guideline (temp = 1) | | |
| --- | --- | --- | --- | --- | --- | --- |
|  | Precision | Recall | F1-Score | Precision | Recall | F1-Score |
| Intracerebral hemorrhage (ICH) | 0.86 (0.65 to 0.95) | 1.00 (0.82 to 1.00) | 0.92 (0.73 to 0.97) | 0.90 (0.70 to 0.97) | 1.00 (0.82 to 1.00) | 0.95 (0.76 to 0.99) |
| Epidural hematoma (EDH) | 0.50 (0.10 to 0.91) | 1.00 (0.21 to 1.00) | 0.67 (0.13 to 0.95) | 0.50 (0.10 to 0.91) | 1.00 (0.21 to 1.00) | 0.67 (0.13 to 0.95) |
| Subdural hematoma (SDH) | 1.00 (0.81 to 1.00) | 1.00 (0.81 to 1.00) | 1.00 (0.81 to 1.00) | 1.00 (0.81 to 1.00) | 1.00 (0.81 to 1.00) | 1.00 (0.81 to 1.00) |
| Subarachnoid hemorrhage (SAH) | 0.94 (0.72 to 0.99) | 0.94 (0.72 to 0.99) | 0.94 (0.72 to 0.99) | 0.94 (0.72 to 0.99) | 0.94 (0.72 to 0.99) | 0.94 (0.72 to 0.99) |
| Infarct demarcation | 0.55 (0.43 to 0.66) | 1.00 (0.90 to 1.00) | 0.71 (0.58 to 0.79) | 0.86 (0.72 to 0.93) | 1.00 (0.90 to 1.00) | 0.92 (0.80 to 0.97) |
| Vascular occlusion | 0.99 (0.94 to 1.00) | 0.99 (0.94 to 1.00) | 0.99 (0.94 to 1.00) | 0.99 (0.94 to 1.00) | 0.98 (0.92 to 0.99) | 0.98 (0.93 to 1.00) |
| Vascular stenosis | 0.90 (0.81 to 0.94) | 1.00 (0.95 to 1.00) | 0.94 (0.88 to 0.97) | 0.96 (0.90 to 0.99) | 1.00 (0.95 to 1.00) | 0.98 (0.92 to 0.99) |
| Aneurysm | 0.90 (0.60 to 0.98) | 1.00 (0.70 to 1.00) | 0.95 (0.64 to 0.99) | 1.00 (0.68 to 1.00) | 0.89 (0.57 to 0.98) | 0.94 (0.62 to 0.99) |
| Dissection | 0.75 (0.30 to 0.95) | 1.00 (0.44 to 1.00) | 0.86 (0.36 to 0.98) | 0.75 (0.30 to 0.95) | 1.00 (0.44 to 1.00) | 0.86 (0.36 to 0.98) |
| Ischemic perfusion deficit | 0.95 (0.89 to 0.98) | 0.98 (0.92 to 0.99) | 0.96 (0.90 to 0.99) | 0.94 (0.88 to 0.98) | 0.99 (0.94 to 1.00) | 0.97 (0.90 to 0.99) |
| Total | 0.87 (0.83 to 0.90) | 0.99 (0.97 to 0.99) | 0.93 (0.90 to 0.94) | 0.95 (0.92 to 0.96) | 0.99 (0.97 to 0.99) | 0.97 (0.94 to 0.98) |

Supplement 1: Data extraction performance of GPT-4o (temperature = 1) with and without annotation guideline in dataset A (n = 200). Metrics for GPT-4o were calculated based on the mode across three repetitions.

| Variable | Dataset A: Llama-3.3-70B – Guideline (temp = 0) | | | Dataset A: Llama-3.3-70B + Guideline (temp = 0) | | |
| --- | --- | --- | --- | --- | --- | --- |
|  | Precision | Recall | F1-Score | Precision | Recall | F1-Score |
| Intracerebral hemorrhage (ICH) | 0.78 (0.58 to 0.90) | 1.00 (0.82 to 1.00) | 0.88 (0.68 to 0.95) | 0.75 (0.55 to 0.88) | 1.00 (0.82 to 1.00) | 0.86 (0.66 to 0.94) |
| Epidural hematoma (EDH) | 0.50 (0.10 to 0.91) | 1.00 (0.21 to 1.00) | 0.67 (0.13 to 0.95) | 0.33 (0.06 to 0.79) | 1.00 (0.21 to 1.00) | 0.50 (0.09 to 0.88) |
| Subdural hematoma (SDH) | 1.00 (0.81 to 1.00) | 1.00 (0.81 to 1.00) | 1.00 (0.81 to 1.00) | 1.00 (0.81 to 1.00) | 1.00 (0.81 to 1.00) | 1.00 (0.81 to 1.00) |
| Subarachnoid hemorrhage (SAH) | 0.94 (0.72 to 0.99) | 0.94 (0.72 to 0.99) | 0.94 (0.72 to 0.99) | 0.94 (0.72 to 0.99) | 0.94 (0.72 to 0.99) | 0.94 (0.72 to 0.99) |
| Infarct demarcation | 0.53 (0.41 to 0.64) | 1.00 (0.90 to 1.00) | 0.69 (0.57 to 0.78) | 0.60 (0.47 to 0.71) | 1.00 (0.90 to 1.00) | 0.75 (0.62 to 0.83) |
| Vascular occlusion | 0.86 (0.78 to 0.91) | 1.00 (0.96 to 1.00) | 0.92 (0.86 to 0.95) | 0.98 (0.92 to 0.99) | 0.98 (0.92 to 0.99) | 0.98 (0.92 to 0.99) |
| Vascular stenosis | 0.67 (0.58 to 0.75) | 1.00 (0.95 to 1.00) | 0.80 (0.72 to 0.86) | 0.86 (0.77 to 0.91) | 1.00 (0.95 to 1.00) | 0.92 (0.85 to 0.95) |
| Aneurysm | 0.90 (0.60 to 0.98) | 1.00 (0.70 to 1.00) | 0.95 (0.64 to 0.99) | 1.00 (0.70 to 1.00) | 1.00 (0.70 to 1.00) | 1.00 (0.70 to 1.00) |
| Dissection | 0.75 (0.30 to 0.95) | 1.00 (0.44 to 1.00) | 0.86 (0.36 to 0.98) | 0.75 (0.30 to 0.95) | 1.00 (0.44 to 1.00) | 0.86 (0.36 to 0.98) |
| Ischemic perfusion deficit | 0.93 (0.86 to 0.97) | 0.99 (0.94 to 1.00) | 0.96 (0.90 to 0.98) | 0.91 (0.84 to 0.95) | 0.99 (0.94 to 1.00) | 0.95 (0.88 to 0.98) |
| Total | 0.78 (0.74 to 0.81) | 0.99 (0.98 to 1.00) | 0.87 (0.84 to 0.90) | 0.86 (0.82 to 0.89) | 0.99 (0.97 to 0.99) | 0.92 (0.89 to 0.94) |

Supplement 2: Data extraction performance of Llama-3.3-70B (temperature = 0) with and without annotation guideline in dataset A (n = 200).

| Variable | Dataset B: Llama-3.3-70B – Guideline (temp = 0) | | | Dataset B: Llama-3.3-70B + Guideline (temp = 0) | | |
| --- | --- | --- | --- | --- | --- | --- |
|  | Precision | Recall | F1-Score | Precision | Recall | F1-Score |
| Intracerebral hemorrhage (ICH) | 0.50 (0.21 to 0.79) | 1.00 (0.51 to 1.00) | 0.67 (0.30 to 0.88) | 0.57 (0.25 to 0.84) | 1.00 (0.51 to 1.00) | 0.73 (0.34 to 0.91) |
| Epidural hematoma (EDH) | 0.00 (0.00 to 0.00) | 0.00 (0.00 to 0.00) | 0.00 (0.00 to 0.00) | 0.00 (0.00 to 0.79) | 0.00 (0.00 to 0.00) | 0.00 (0.00 to 0.00) |
| Subdural hematoma (SDH) | 0.67 (0.21 to 0.94) | 1.00 (0.34 to 1.00) | 0.80 (0.26 to 0.97) | 1.00 (0.34 to 1.00) | 1.00 (0.34 to 1.00) | 1.00 (0.34 to 1.00) |
| Subarachnoid hemorrhage (SAH) | 0.50 (0.10 to 0.91) | 1.00 (0.21 to 1.00) | 0.67 (0.13 to 0.95) | 0.50 (0.10 to 0.91) | 1.00 (0.21 to 1.00) | 0.67 (0.13 to 0.95) |
| Infarct demarcation | 0.50 (0.31 to 0.69) | 1.00 (0.74 to 1.00) | 0.67 (0.43 to 0.82) | 0.58 (0.36 to 0.77) | 1.00 (0.74 to 1.00) | 0.73 (0.49 to 0.87) |
| Vascular occlusion | 0.84 (0.65 to 0.94) | 1.00 (0.84 to 1.00) | 0.91 (0.74 to 0.97) | 0.95 (0.78 to 0.99) | 1.00 (0.84 to 1.00) | 0.98 (0.81 to 1.00) |
| Vascular stenosis | 0.53 (0.39 to 0.66) | 1.00 (0.87 to 1.00) | 0.69 (0.54 to 0.80) | 0.67 (0.51 to 0.79) | 1.00 (0.87 to 1.00) | 0.80 (0.64 to 0.89) |
| Aneurysm | 0.67 (0.30 to 0.90) | 1.00 (0.51 to 1.00) | 0.80 (0.38 to 0.95) | 0.80 (0.38 to 0.96) | 1.00 (0.51 to 1.00) | 0.89 (0.43 to 0.98) |
| Dissection | 0.50 (0.10 to 0.91) | 1.00 (0.21 to 1.00) | 0.67 (0.13 to 0.95) | 0.50 (0.10 to 0.91) | 1.00 (0.21 to 1.00) | 0.67 (0.13 to 0.95) |
| Ischemic perfusion deficit | 0.86 (0.69 to 0.94) | 0.96 (0.81 to 0.99) | 0.91 (0.74 to 0.97) | 0.85 (0.68 to 0.94) | 0.92 (0.75 to 0.98) | 0.89 (0.71 to 0.96) |
| Total | 0.65 (0.57 to 0.72) | 0.99 (0.94 to 1.00) | 0.78 (0.71 to 0.84) | 0.74 (0.66 to 0.81) | 0.98 (0.93 to 0.99) | 0.84 (0.77 to 0.89) |

Supplement 3: Data extraction performance of Llama-3.3-70B (temperature = 0) with and without annotation guideline in dataset B (n = 100).

| Variable | Dataset A: GPT-4o – Guideline (temp = 0) | | | Dataset A: GPT-4o + Guideline (temp = 0) | | |
| --- | --- | --- | --- | --- | --- | --- |
|  | Precision | Recall | F1-Score | Precision | Recall | F1-Score |
| Intracerebral hemorrhage (ICH) | 0.86 (0.65 to 0.95) | 1.00 (0.82 to 1.00) | 0.92 (0.73 to 0.97) | 0.90 (0.70 to 0.97) | 1.00 (0.82 to 1.00) | 0.95 (0.76 to 0.99) |
| Epidural hematoma (EDH) | 0.50 (0.10 to 0.91) | 1.00 (0.21 to 1.00) | 0.67 (0.13 to 0.95) | 0.50 (0.10 to 0.91) | 1.00 (0.21 to 1.00) | 0.67 (0.13 to 0.95) |
| Subdural hematoma (SDH) | 1.00 (0.81 to 1.00) | 1.00 (0.81 to 1.00) | 1.00 (0.81 to 1.00) | 1.00 (0.81 to 1.00) | 1.00 (0.81 to 1.00) | 1.00 (0.81 to 1.00) |
| Subarachnoid hemorrhage (SAH) | 0.94 (0.72 to 0.99) | 0.94 (0.72 to 0.99) | 0.94 (0.72 to 0.99) | 1.00 (0.80 to 1.00) | 0.94 (0.72 to 0.99) | 0.97 (0.75 to 0.99) |
| Infarct demarcation | 0.52 (0.41 to 0.64) | 1.00 (0.90 to 1.00) | 0.69 (0.56 to 0.78) | 0.84 (0.70 to 0.92) | 1.00 (0.90 to 1.00) | 0.91 (0.79 to 0.96) |
| Vascular occlusion | 0.99 (0.94 to 1.00) | 0.99 (0.94 to 1.00) | 0.99 (0.94 to 1.00) | 0.99 (0.94 to 1.00) | 0.99 (0.94 to 1.00) | 0.99 (0.94 to 1.00) |
| Vascular stenosis | 0.89 (0.80 to 0.94) | 1.00 (0.95 to 1.00) | 0.94 (0.87 to 0.97) | 0.97 (0.91 to 0.99) | 0.96 (0.89 to 0.99) | 0.97 (0.90 to 0.99) |
| Aneurysm | 0.90 (0.60 to 0.98) | 1.00 (0.70 to 1.00) | 0.95 (0.64 to 0.99) | 1.00 (0.70 to 1.00) | 1.00 (0.70 to 1.00) | 1.00 (0.70 to 1.00) |
| Dissection | 0.75 (0.30 to 0.95) | 1.00 (0.44 to 1.00) | 0.86 (0.36 to 0.98) | 0.75 (0.30 to 0.95) | 1.00 (0.44 to 1.00) | 0.86 (0.36 to 0.98) |
| Ischemic perfusion deficit | 0.94 (0.87 to 0.98) | 0.98 (0.92 to 0.99) | 0.96 (0.90 to 0.98) | 0.94 (0.88 to 0.98) | 1.00 (0.96 to 1.00) | 0.97 (0.92 to 0.99) |
| Total | 0.86 (0.82 to 0.89) | 0.99 (0.97 to 1.00) | 0.92 (0.89 to 0.94) | 0.95 (0.92 to 0.97) | 0.99 (0.97 to 0.99) | 0.97 (0.94 to 0.98) |

Supplement 4: Data extraction performance of GPT-4o (temperature = 0) with and without annotation guideline in dataset A (n = 100).

| Variable | Dataset B: GPT-4o – Guideline (temp = 0) | | | Dataset B: GPT-4o + Guideline (temp = 0) | | |
| --- | --- | --- | --- | --- | --- | --- |
|  | Precision | Recall | F1-Score | Precision | Recall | F1-Score |
| Intracerebral hemorrhage (ICH) | 1.00 (0.51 to 1.00) | 1.00 (0.51 to 1.00) | 1.00 (0.51 to 1.00) | 1.00  (0.51 to 1.00) | 1.00  (0.51 to 1.00) | 1.00  (0.51 to 1.00) |
| Epidural hematoma (EDH) | 0.00 (0.00 to 0.00) | 0.00 (0.00 to 0.00) | 0.00 (0.00 to 0.00) | 0.00  (0.00 to 0.00) | 0.00  (0.00 to 0.00) | 0.00  (0.00 to 0.00) |
| Subdural hematoma (SDH) | 0.67 (0.21 to 0.94) | 1.00 (0.34 to 1.00) | 0.80 (0.26 to 0.97) | 1.00  (0.34 to 1.00) | 1.00  (0.34 to 1.00) | 1.00  (0.34 to 1.00) |
| Subarachnoid hemorrhage (SAH) | 0.50 (0.10 to 0.91) | 1.00 (0.21 to 1.00) | 0.67 (0.13 to 0.95) | 0.50  (0.10 to 0.91) | 1.00  (0.21 to 1.00) | 0.67  (0.13 to 0.95) |
| Infarct demarcation | 0.65 (0.41 to 0.83) | 1.00 (0.74 to 1.00) | 0.79 (0.53 to 0.91) | 0.85  (0.58 to 0.96) | 1.00  (0.74 to 1.00) | 0.92  (0.65 to 0.98) |
| Vascular occlusion | 1.00 (0.85 to 1.00) | 1.00 (0.85 to 1.00) | 1.00 (0.85 to 1.00) | 1.00  (0.85 to 1.00) | 1.00  (0.85 to 1.00) | 1.00  (0.85 to 1.00) |
| Vascular stenosis | 0.84 (0.67 to 0.93) | 1.00 (0.87 to 1.00) | 0.91 (0.76 to 0.96) | 0.96  (0.81 to 0.99) | 0.96  (0.81 to 0.99) | 0.96  (0.81 to 0.99) |
| Aneurysm | 0.67 (0.30 to 0.90) | 1.00 (0.51 to 1.00) | 0.80 (0.38 to 0.95) | 0.80  (0.38 to 0.96) | 1.00  (0.51 to 1.00) | 0.89  (0.43 to 0.98) |
| Dissection | 0.50 (0.10 to 0.91) | 1.00 (0.21 to 1.00) | 0.67 (0.13 to 0.95) | 0.50  (0.10 to 0.91) | 1.00  (0.21 to 1.00) | 0.67  (0.13 to 0.95) |
| Ischemic perfusion deficit | 1.00 (0.86 to 1.00) | 0.96 (0.81 to 0.99) | 0.98 (0.83 to 1.00) | 0.96  (0.81 to 0.99) | 0.96  (0.81 to 0.99) | 0.96  (0.81 to 0.99) |
| Total | 0.86 (0.78 to 0.91) | 0.99 (0.94 to 1.00) | 0.92 (0.85 to 0.95) | 0.93  (0.86 to 0.97) | 0.98  (0.93 to 0.99) | 0.95  (0.89 to 0.98) |

Supplement 5: Data extraction performance of GPT-4o (temperature = 0) with and without annotation guideline in dataset B (n = 100).
